# Probiotics-induced changes in gut microbial composition and its effects on cognitive performance after stress

**DOI:** 10.1101/2020.11.23.20236836

**Authors:** Mirjam Bloemendaal, Joanna Szopinska-Tokov, Clara Belzer, David Boverhoff, Silvia Papalini, Franziska Michels, Saskia van Hemert, Alejandro Arias Vasquez, Esther Aarts

## Abstract

Stress negatively affects cognitive performance. Probiotics remediate somatic and behavioral stress responses, hypothetically by acting on the gut microbiota. Here, we assessed gut microbial alterations after 28-days supplementation of multi-strain probiotics (EcologicBarrier consisting of *Lactobacilli, Lactococci* and *Bifidobacteria* in healthy, female subjects (probiotics group n=27, placebo group n=29). In an identical pre- and post-session, subjects performed a working memory task before and after an acute stress intervention. One-month supplementation of probiotics changed the gut microbial community (within probiotics group P_weighted unifrac_=0.008). This change was driven by a nominal increase in eight genera in the probiotics group relative to the placebo group: *Butyricimonas, Parabacteroides, Alistipes, Christensenellaceae_R-7_group, Family_XIII_AD3011_group, Ruminococcaceae_UCG-003, Ruminococcaceae_UCG-005* and *Ruminococcaceae_UCG-010*. Of these, the probiotics-induced change in genus *Ruminococcaceae_UCG-003* was significantly associated with probiotics’ effect on stress-induced working memory changes (r_spearman_(27) = 0.565; pFDR = 0.014) in the probiotics group only and independent of potential confounders (i.e., age, BMI, and baseline dietary fibre intake). That is, subjects with a higher increase in *Ruminococcaceae_UCG-003* abundance after probiotics were also more protected from negative effects of stress on working memory after probiotic supplementation. The bacterial taxa showing an increase in relative abundance in the probiotics group are plant fibre degrading bacteria and produce short-chain fatty acids that are known for their beneficial effect on gut and brain health, e.g. maintaining intestinal- and blood-brain-barrier integrity. This study shows that gut microbial alterations, modulated through probiotics use, are related to improved cognitive performance in acute stress circumstances.

## Introduction

Stress, regardless of its origin (physical, mental or social), activates the hypothalamic-pituitary-adrenal (HPA) axis ^1^; a neuroendocrine system that controls the body’s stress response. Stress-induced glucocorticoids released by the adrenal cortex impact many tissues in the body, including the brain where it affects cognitive performance (e.g., working memory ^2^). Moreover, stress plays an important role in the neurobiology of mood disorders, including depressive and bipolar disorders ^3^. The systemic effects of stress includes mediation by the gut-brain axis (GBA) (for a review see Foster et al. ^4^). This axis refers to the bidirectional communication between the gastrointestinal tract and the central nervous system, which is not only mediated by endocrine signaling including hormones and other neuro-active metabolites, but also by the vagus nerve and by the immune system ^5^. The gut bacteria (gut microbiota) modify the functioning of the GBA ^6^, making it a key player in behavior and stress reactivity ^4^. A strong illustration of the role of the microbiota in the GBA and its effect on (healthy) behavior are the results of the experiments carried out in germ-free mice (i.e., mice reared in a germ-free environment preventing colonization of the intestinal tract) or animals subjected to wash out of their bacterial community by antibiotics. These animal models clearly show cognitive impairments, multiple behavioral disturbances such as altered anxiety responses, learning problems and exaggerated stress reactivity ^7,8^. Sudo et al. ^9^ observed altered HPA axis functioning in germ-free mice, i.e., amplified corticosterone response to stress induced by physical restraint, compared with control mice.

The gut microbiota are a promising target for protection against negative effects of stress, as it is modifiable through e.g. diet or probiotics ^4^. Probiotics are defined as “live, micro-organisms, which induces a health benefit to the host when administered in adequate amounts” ^10^. Use of probiotics can be advised in case of irregularities in digestion and or stool composition such as diarrhea, aiming to re-establish more diverse gut microbiota abundance and reduce gastrointestinal complaints ^11^. With the increasing awareness of the importance of the link between the microbiota-GBA in mood and stress-related symptoms, the potential of probiotics in protecting and or restoring these symptoms has been explored ^12,13^.

In the above-mentioned experiment in germ-free mice, Sudo et al. ^9^ showed that the amplified corticosterone response to stress was normalized after supplementation with the bacterial strain *Bifidobacterium infantis*. Other animal studies showed that probiotics reduces anxious behaviors in response to physical stressors: i.e., less defensive probe burying after receiving shocks when supplemented with *Lactobacillus helveticus R0052* and *Bifidobacterium longum R0175* for two-weeks ^14^; more time spent in an open field after hypothermia when supplemented with *L. rhamnosus* for 29 days ^15^; and reduced immobilization time in a forced swim test after 5-week supplementation of the same multi-species probiotics mix as used in the current study ^16^.

In human randomized placebo-controlled trials (RCTs), probiotics have also demonstrated beneficial effects on physical and/or psychological complaints in response to experimental acute stress paradigms or daily life stress. For example, subjects showed reduced depressive symptoms after 30-days supplementation of the same 2-strain probiotic product effective in the animal study mentioned above ^14^. Adults experiencing moderate stress symptoms on the Perceived Stress Scale (PSS-10) at baseline reported fewer of these symptoms after 8-weeks use of *Lactobaccilus Plantarum* DR7 strain ^17^ or after 12-weeks use of the P8 strain of the same species ^18^. Reduced abdominal complaints during exam stress were observed using *Lactobacillus casei* for 8 weeks ^19^. Similarly, eight-weeks supplementation of the *Lactobacillus casei* strain Shirota YIT 9029 relieved abdominal pain and exam stress ^20^. After an acute stress paradigm, i.e. the socially evaluated cold pressor test (SECPT), fewer self-reported anxiety symptoms were observed after 4 weeks of *Bifidobacterium longum* 1714 use than after placebo ^21^. In contrast, in two other RTCs in healthy volunteers, no alterations in stress-related measures were observed after two-weeks supplementation of *Lactobacilli, Bifidobacteria* and *Streptococci* strains ^22^ or after eight-week *Lactobacillus rhamnosus* supplementation ^23^. Thus, although probiotics have been shown to affect human stress-related measures in some studies, the effects are not always replicated and more research is needed to understand the mechanisms behind these effects.

The negative effects of (acute) stress on cognitive functioning have been rarely assessed in a probiotics trial. The double-blind RCT reporting fewer stress symptoms after 8-weeks *Lactobaccilus Plantarum* DR7 strain exposure also observed probiotic-induced improvements in emotion recognition speed and verbal memory speed ^17^. Allen et al. observed fewer stress (SECPT) induced errors on a visuospatial memory task after 4 weeks of *Bifidobacterium longum* 1714 ^21^. Moreover, Papalini et al. ^24^ assessed the effect of 4-week multi-strain probiotics supplementation on neuro-cognitive functioning in a double-blind RTC in healthy female volunteers. Probiotics protected against working memory detriments caused by an acute stress paradigm. That is, the probiotics group performed better after (versus before) the SECPT on a digit span backward test compared with the placebo group. Furthermore, this protecting effect of probiotics on working-memory decline after stress was associated with changes in prefrontal cortex fMRI signal during cognitive control.

From the above studies it is, however, unclear how probiotics could exert their effect on central neural processing and cognitive performance. One mechanism underlying the effects of probiotics on the GBA could be a specific alteration in microbial composition. Measuring probiotics-induced gut microbial alterations can help identify which bacteria mediate the protecting effects of probiotics on mental health and cognitive performance and can provide mechanistic insights into e.g. metabolite production and other microbial functions ^25^. Yet, almost none of the above-mentioned studies report the effect of probiotics supplementation on gut microbial profiling. Kato-Kataoka et al. ^20^ did observe higher alpha gut microbial diversity in the probiotics group in addition to lowered stress symptoms. However, the authors did not report an association between these measures of stress and microbiota. Another microbiota-GBA study assessed the effects of a *Lactobacilli* and *Lactococci* probiotics mixture on the gut microbial composition and emotional memory of healthy volunteers and found that increased memory was associated with decreased abundance of nine genera ^26^. However, these results were not observed in the context of stress-reactivity.

Here, we aimed to assess how probiotics can buffer against the detrimental effects of stress on cognition by investigating the link with the probiotics-induced changes in the gut microbiota. For this, we used the data of the Papalini et al. ^24^ study, where 28-days multi-species probiotics versus placebo supplementation was shown to improve working memory (i.e. digit span backwards) performance after an acute stressor (i.e. the socially evaluated cold pressor test) in healthy female volunteers. We extend this work by assessing, first, whether this probiotics product alters gut microbial composition versus placebo and, second, whether this probiotics’ effect on gut microbial composition relates to the probiotics’ protective effect on stress-induced working memory changes.

## Methods

### Subjects

Healthy female subjects, aged between 18 and 40 years old were included (see **Supplementary Materials** for further inclusion criteria). A total of 61 healthy subjects were tested, of which one subject was excluded from the study due to high scores on the Beck Depression Inventory questionnaire (BDI), potentially indicating depression. For two subjects the pre-intervention fecal material was lacking, resulting in 58 subjects for the gut microbiota analysis (n=27 in the probiotics group, n=31 in the placebo group) (see **Supplementary Figure 1**). Two other subjects scored poorly on the behavioral tasks and were excluded, resulting in a total sample - including both microbiota and neuro-cognitive data - of 56 subjects (probiotics group: n = 27, mean age = 21.8, SEM = 0.5, mean BMI = 21.9, SEM= 0.32; placebo group: n = 29, mean age = 22.4, SEM = 0.53, mean BMI = 21.7, SEM= 0.30). The groups did not differ in age, BMI and neither in baseline dietary intake as reported in the Food Frequency Questionnaire Dutch Healthy Diet (FFQ-DHD) ^27^, see **Supplementary Table 1**.

The study was conducted following the Declaration of Helsinki with human subjects and the complete procedure was approved by the local Ethics Committee (CMO Arnhem-Nijmegen, NL55406.091.15) and registered at the Dutch trial register (protocol number: NTR5845). Written informed consent was obtained from each participant. For an exhaustive description of the methods in this study, see Papalini et al.^24^, of which this study is an extension.

### Procedure

Subjects were tested on two days: the first day (baseline), before the intervention started, and the second day after 28 days (four weeks) of probiotics or placebo administration. The identical test sessions included cognitive testing in and outside the MRI scanner, an acute stress intervention, and fecal material collection before starting and after the last day of the intervention (see **Figure 1**).

**Figure 1.**
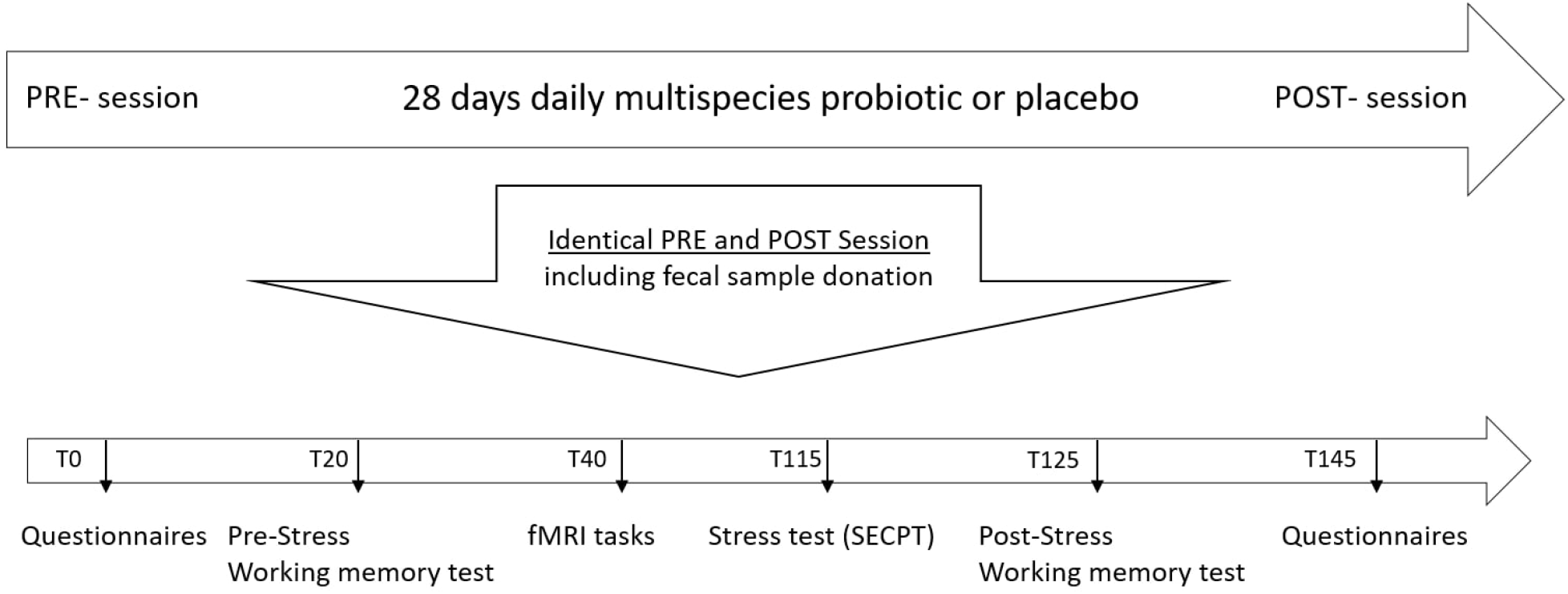
Overview of the testing sessions. Each participant was tested twice, before and after 4 weeks of supplementation with probiotics/placebo. The procedure of the two sessions was the same (i.e. subjects performed the same tests in the same order). Tx: x minutes since the start of the test session. SECPT: socially evaluated cold pressor test.

Relevant for the current analyses, the working memory task (digit span forward and backward) was performed before and after the SECPT. The SECPT is an established stress-inducing paradigm consisting of both a physical and a social stressor ^28^. Also in this experiment, the SECPT increased subjective feelings of stress measured with Visual Analogue Scales (VAS) and increased physiological measures, i.e. heart rate, blood pressure and cortisol levels, in our subjects (see Figure 4 in Papalini et al.^24^). During the digit span task, subjects listen to a series of numbers and are instructed to repeat each series correctly (digit span forward) or repeat it backwards (digit span backward). Following a correct response, increasingly longer sequences are presented to the participant. Different series of numbers are used on each occasion (before and after stress on the first and second test day) to avoid long-term memory effects. In the SECPT ^28^, subjects were instructed to hold their hand in a bucket of ice water (0 – 3°C) for as long as possible (limited to three minutes) under surveillance of a video camera and an unfamiliar, disesteem expressing researcher. After the SECPT, the digit span was re-tested, hereby enabling assessment of stress on working memory performance. As in our previous work ^24^, we only focus on the digit span backward, given that SECPT specifically influenced this type of working memory modulation instead of simply working memory maintenance that is needed in the digit span forward ^29^.

Throughout the test session, VAS were used to assess well-being (5 times), heart rate and blood pressure was measured (7 times), and saliva samples (5 times) were obtained. Three fMRI paradigms measuring different aspects of emotion and cognition were performed while scanning. Once per test session, we further evaluated mood, emotional state, sensitivity, and diet. The total test session lasted almost three hours. The effects of the probiotics versus placebo intervention on these measures have already been reported in Papalini et al. ^24^.

### Intervention

We assessed the GBA mechanisms of a probiotics product called Ecologic®Barrier (Winclove, The Netherlands). This product consists of the following bacterial strains: *Bifidobacterium bifidum* W23, *Bifidobacterium lactis* W51, *Bifidobacterium lactis* W52, *Lactobacillus acidophilus* W37, *Lactobacillus brevis* W63, *Lactobacillus casei* W56, *Lactobacillus salivarius* W24, *Lactococcus lactis* W19 and *Lactococcus lactis* W58. The total cell count was 2.5 x 10^9 colony forming units (cfu) per gram, and subjects consumed 2 grams per day i.e. 5 x 10^9 cfu per day. More information on the product in the **Supplementary Materials**.

### Microbiome sample processing

#### Fecal sample collection

Fecal samples were collected by the subjects at home using a validated protocol by OMNIgene•GUT kit (DNAGenotek, Ottawa, CA). The material was aliquoted into 1.5 ml Eppendorf tubes and stored in -80 °C until further laboratory processing.

#### Bacterial DNA isolation and sequencing

For bacterial DNA extraction, 50 mg of faeces was separated from 200 µL of OMNIgene•GUT kit buffer by centrifugation at 1400 rcf at 16°C for 5min. Microbial DNA was isolated from faecal pellets using the Maxwell® 16 Instrument (Promega, Leiden, The Netherlands) as described previously ^2^. DNA purification was performed with a customized kit (AS1220; Promega) using 250 μl of the final supernatant pool. The V4 region of 16S ribosomal RNA (rRNA) gene was amplified in duplicate PCR reactions for each sample in a total reaction volume of 50 μl. The V4 region was targeted by using previously reported primers for this region: 515F (5’GTGYCAGCMGCCGCGGTAA) – 806R (5’ GGACTACNVGGGTWTCTAAT) ^2^. We included a PCR negative sample to assess contamination introduced during this step. The purified samples were used to prepare libraries for the Illumina HiSeq PE300 (paired-end, 300 bp) sequencing platform (GATC Biotech AG, Konstanz, Germany), with final loading concentrations of 200 ng/µ. For more information on the wet lab procedure, see ^3^.

### Data analyses

#### Bioinformatics

Using the NG-Tax 16S rRNA pipeline ^32^, taxonomic information was assigned. Briefly, the pipeline can be defined by three core elements: (i) barcode-primer filtering, (ii) operational taxonomic unit (OTU) picking and (iii) taxonomic assignment using the SILVA reference database (version 128). Two filtering steps were applied to the output file (BIOM-file) of NG-Tax. First, a genus was filtered based on 10% prevalence cutoff. Second, a sample with less than 10% of genera was removed (see ^31^). To avoid bias in assessing the effects of the intervention, filtering was done on the pre-intervention session. In total, 9,819,945 high-quality sequences were obtained from all samples after NG-Tax pipeline. These sequences were represented by 1043 OTUs and 175 genera. After the filtering steps, we obtained 9,681,326 sequences represented by 898 OTUs and 86 genera. No samples were removed during these steps.

### Community analyses

#### Alpha-diversity

Alpha-diversity was estimated in three ways using QIIME 1.9.1 ^34^: i) a richness measure counting observed OTUs, ii) Shannon Index taking into account also abundance of the counted OTU’s and iii) Faith’s Phylogenetic Diversity, taking into account the evolutionary phylogenetic structure. After normality was verified, repeated measures ANOVAs were performed on between-subjects factor Intervention (Placebo, Probiotics) and within-subjects factor Time (Pre, Post), assessing Time (Pre, Post) * Intervention (Placebo, Probiotics) as well as a-priori hypothesized simple effects. The results are presented as differences (delta) between the pre- and post-intervention (after 4 weeks), symbolized by Δ.

#### Beta-diversity

The beta-diversity was calculated based on weighted UniFrac ^35^. Weighted UniFrac is a distance metric; explaining differences in the relative abundances between microbial communities based on their evolutionary phylogenetic structure. Permutation testing was performed using the R function vegan::adonis ^36^, testing for interaction effects of Time (Pre, Post) * Intervention (Placebo, Probiotics) as well as a-priori hypothesized simple effects in the weighted UniFrac distance metric, accounting for repeated measures by using the ‘strata’ argument when testing effects of time. To visualize intervention effect on beta-diversity Constrained Analysis of Principal Coordinates (CAP) was performed using the R function vegan::capscale ^37^.

### Composition analyses

#### Taxonomic Differences

The differences in microbial communities between pre- and post-intervention groups were determined at the phylum and genus levels, based on the bacterial relative abundance profiles. Significant differences in relative abundance difference values, post-versus pre-intervention (symbolized by Δ), between probiotics and placebo groups were identified by non-parametric Mann-Whitney U tests. Genera showing an effect of the probiotics intervention were selected for further association analyses with neuro-cognitive measures. Of these, genera with relative abundance levels differing between groups at baseline were not further analysed (see **Figure 2**).

**Figure 2.**
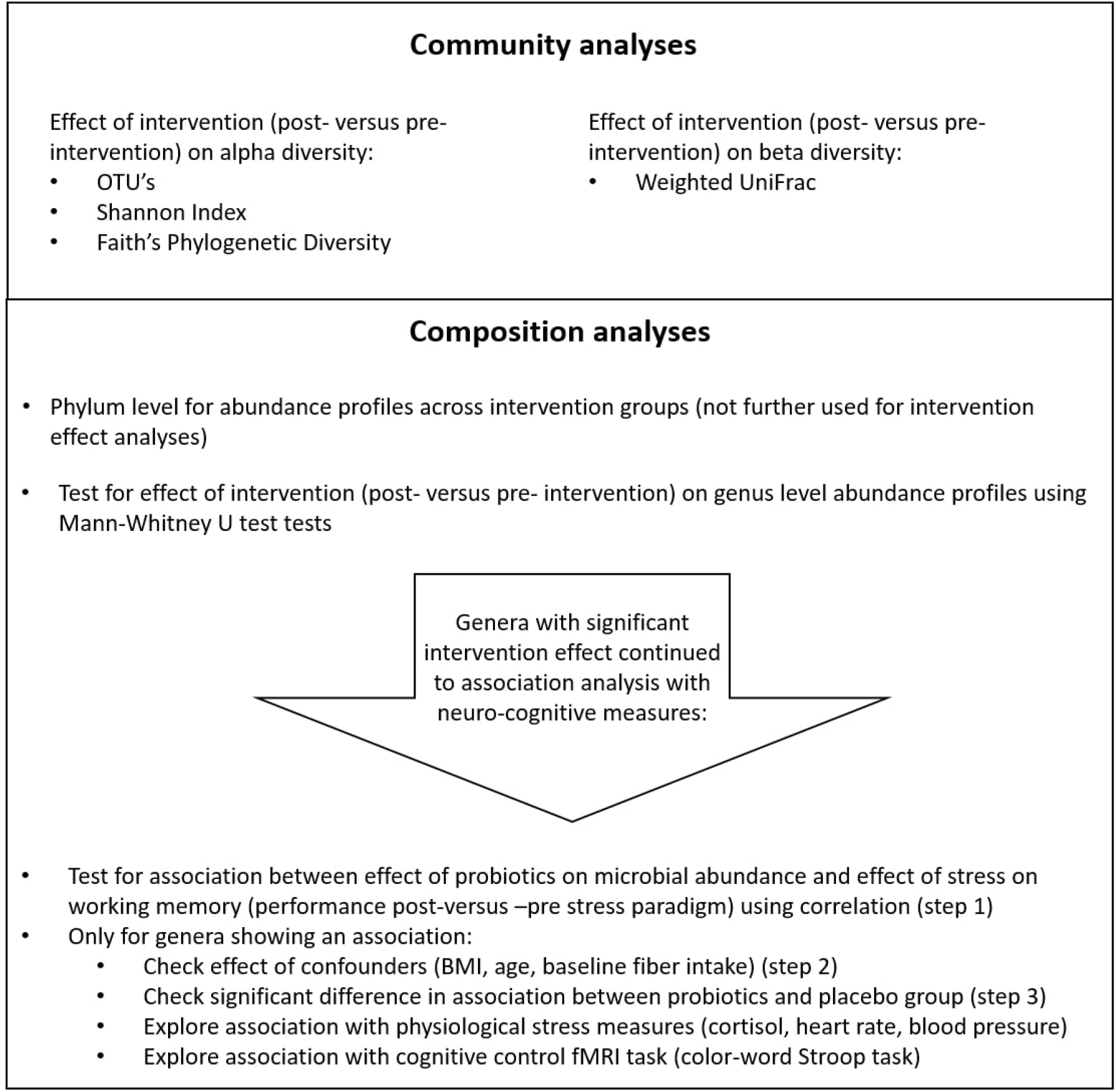
Analysis flow chart. Community dynamics were compared between intervention (post-versus pre-intervention) groups. The composition of the two intervention groups was compared at genus level and related to neuro-cognitive measures.

### Association analyses between intervention-induced changes in the gut microbiota and neuro-cognition

As reported in Papalini et al., 2018, after one-month supplementation of probiotics, the scores of the digit span backward improved after the stress-inducing SECPT paradigm, while this effect was not observed in the placebo group. The association analysis focused primarily on this result: assessing whether the protective effect of probiotics on stress-induced working memory changes is explained by changes in gut microbial composition due to probiotics use. To this aim, firstly, only genera altered due to probiotics exposure (i.e. post-pre increase or decrease (Δ) in relative abundance after probiotics versus placebo) were selected for the downstream analysis. Using correlation analyses, these genera were screened for a relation with the effect of probiotics (post – pre) on working memory (digit span backward, DS) after versus before stress, i.e. (DSafter - DSbefore stress)^post-intervention^ - (DSafter - DSbefore stress)^pre-intervention^ (see **Figure 2**).

Due to the overall skewness of the microbiota data, two-tailed non-parametric Spearman correlations were performed. Only results passing False Discovery Rate (FDR) correction were analysed in a multiple regression analysis. Here, the effect of potential confounders was assessed by adding BMI, age and baseline fibre intake (measured by the DHD) in a linear regression model. Efficacy of probiotics may vary with age and BMI due to metabolic and gastro-intestinal differences ^38,39^. Although no changes in dietary pattern were observed over the course of the intervention, baseline fibre intake may still be a modulator of the effect of probiotics as these serve as a nutrient source for the microbiota ^40^.

The specificity of the association between the effect of probiotics on gut microbial abundance and stress-related working memory was assessed by comparing the regression slopes between the probiotics and placebo group. This was done by creating a dummy variable that divided the intervention groups and subsequently creating an interaction term between this dummy variable and the genus variable. This interaction term was then added in a new regression, assessing whether the relative abundance of the selected genus differentially affects stress-induced working memory changes in the two intervention groups.

For exploratory analyses were performed on fMRI signal and physiological stress measures, see **Supplementary Methods**. The statistical analyses were performed using IBM SPSS statistics (package version 23).

## Results

### Gut microbiota composition before intervention

The intestinal microbiota of the 58 subjects before the intervention (Pre) was typical for healthy individuals; dominated largely by the phyla Firmicutes (68.0%) and Bacteroidetes (19.5%), accounting for up to 87.5% of the intestinal microbial communities. Other observed phyla were Actinobacteria (8.7%), Proteobacteria (1.5%), Verrucomicrobia (1.4%), Euryarchaeota (0.4%), Tenericutes (0.29%) and Cyanobacteria (0.25%).

### Effect of probiotics on gut microbiota community

The three alpha diversity measures did not show significant interaction effects between Time and Intervention, or simple effects in the probiotics and placebo groups, all p>0.05 (**Supplementary Figure 2**).

Beta-diversity at the operational taxonomic unit (OTU) level did not show a significant interaction between Time (pre, post) and treatment (placebo, probiotics) (p=0.244). However, there was a main effect of Time across Interventions (p=0.011). Planned comparisons show that the Time effect was significant within the probiotics group; post versus pre 4-week of intervention (p=0.008), but not in the placebo group (p =0.21) (**Supplementary Table 2**). That is, the post-probiotics samples were more similar among each other compared with the pre-probiotics samples. Constrained analysis of principle coordinates (CAP) ordination method was used to visualize this effect (accounting for 1.3% variability in the dataset) (**Figure 3**).

**Figure 3.**
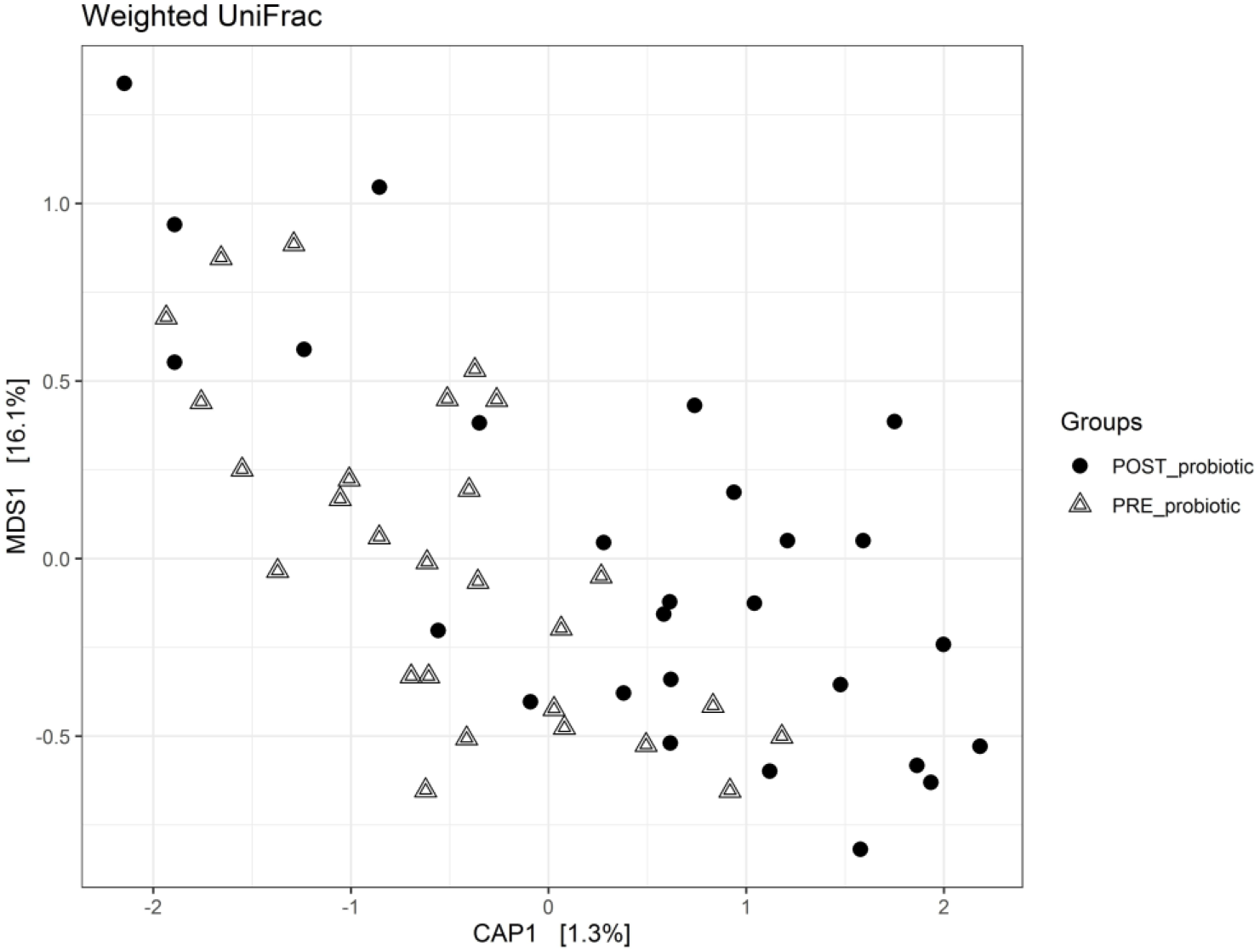
Beta diversity (weigthed Unifrac dissimilarity matrix) in the probiotics group plotted using Canonical Analysis of Principal coordinates (CAP) ordination.

### Effect of probiotics on gut microbiota composition

At the phylum level, we did not find significant differences between a post-pre increase or decrease (Δ) in relative abundance after probiotics versus placebo; **Supplementary Table 3**). At the genus level, we identified nine genera with FDR-uncorrected significant changes in relative abundance in the probiotics group (post-vs. pre-intervention). One of these nine (*Parabacteroides*) was different at baseline between the probiotics and placebo group and was therefore not taken into consideration for further analyses (**Table 1, Supplementary Figure 3A-H**). Another classification (*Lachnospiraceae_g_)* is in fact a sequence for which no classification to an OTU could be made with sufficient confidence, resulting in a grouping of OTU’s with insufficient confidence. This grouping of genera within the *Lachnospiraceae* family was also not taken forward for further analysis.

**Table 1.**
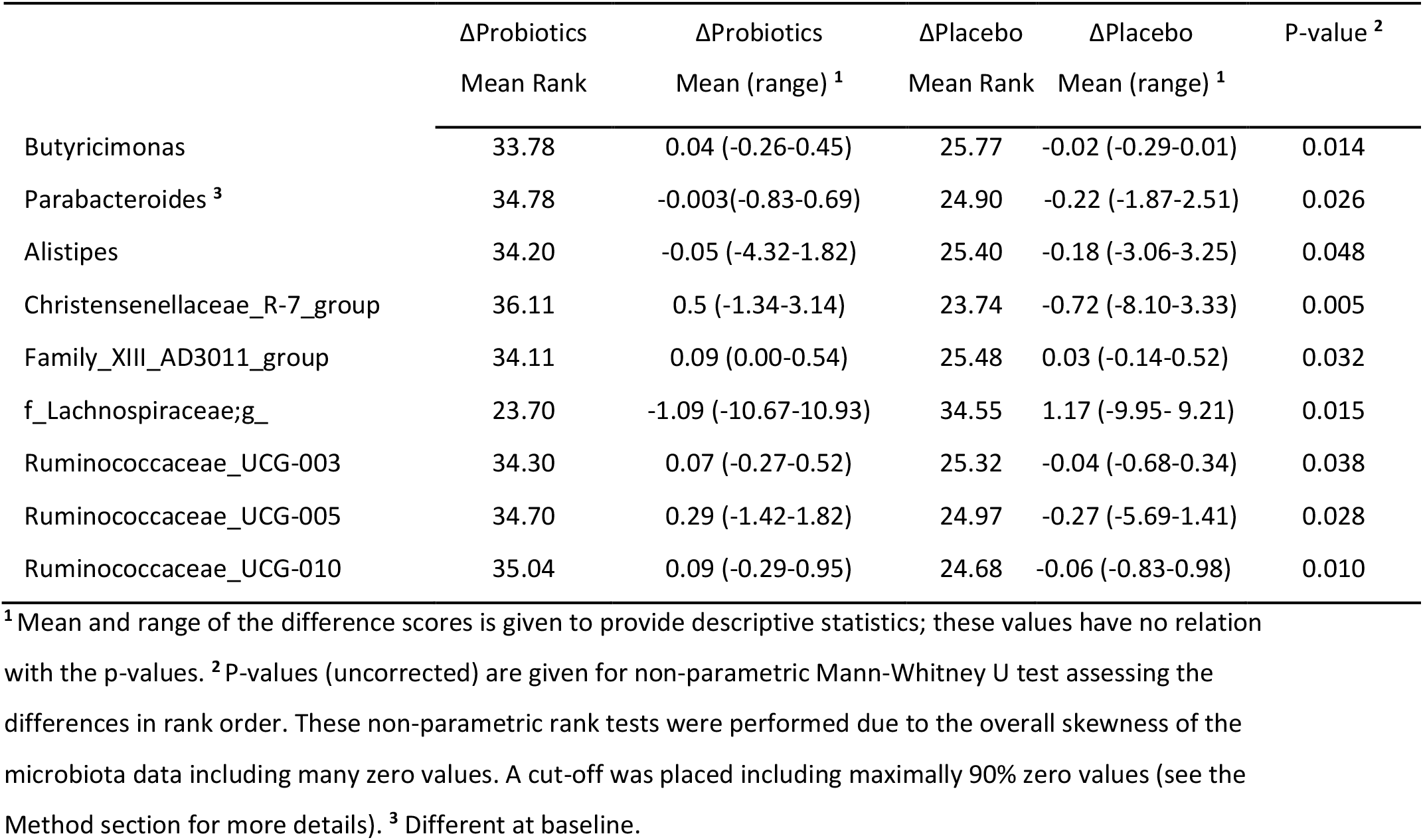
Comparison of genus relative abundance post-pre intervention changes (Δ) between probiotics and placebo groups.

### Association between gut microbiota composition and neurocognitive measures

Seven genera were used to assess a correlation with the protective effect of probiotics on working memory (digit span backward scores) after stress ^24^. Of these, the post-versus pre-change in the relative abundance of genus *Ruminococcaceae_UCG-003* was significantly correlated with the change in the effect of stress on working memory in the probiotics group (r_spearman_(27) = 0.565; pFDR = 0.014) (see **Figure 4**). In the probiotics group, a larger increase (post-versus pre intervention) in the relative abundance of *Ruminococcaceae_UCG-003* was associated with higher (post-versus pre intervention) stress-related working memory performance (digit span backward, post-versus pre-SECPT).

**Figure 4.**
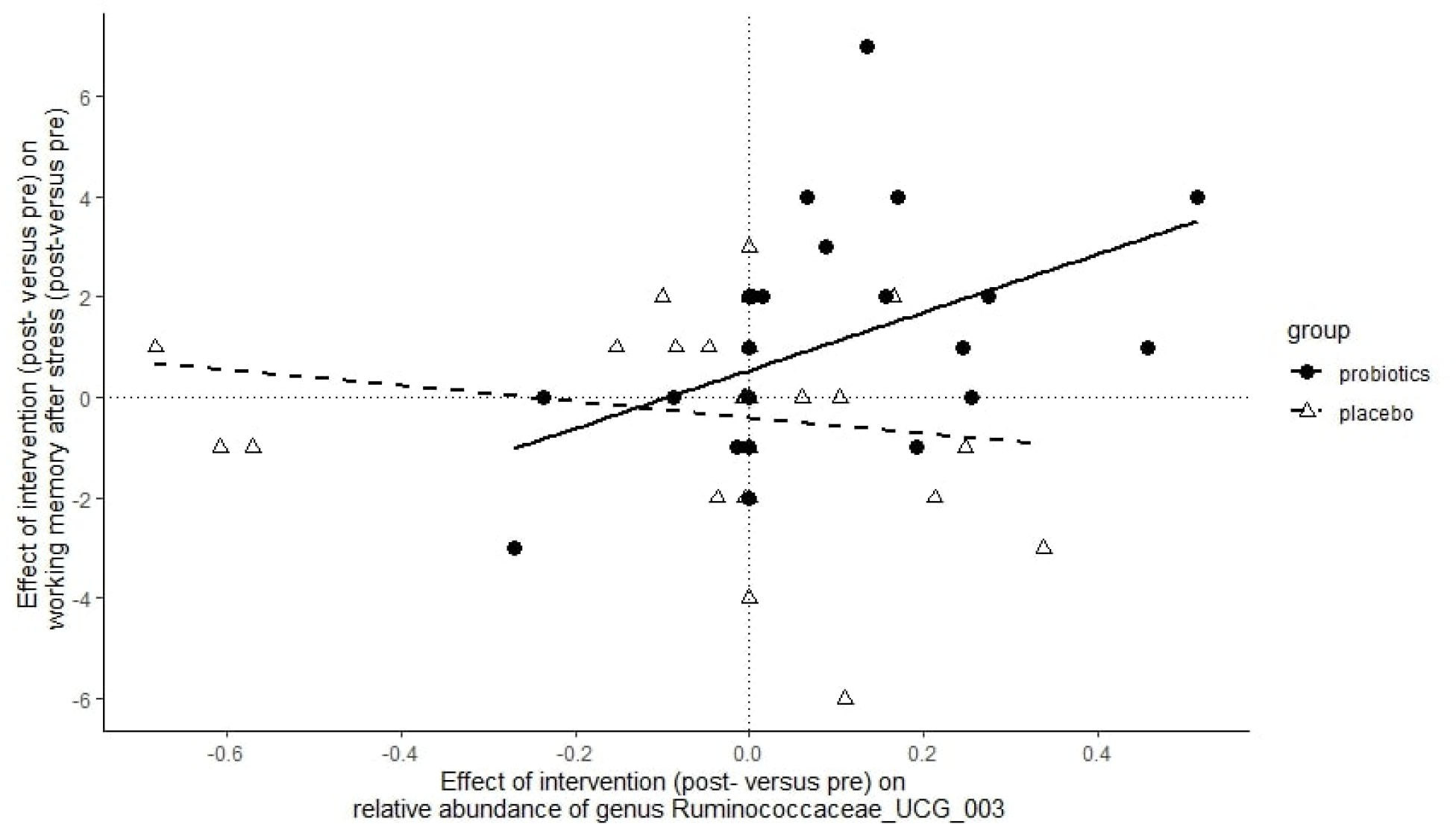
Association between the intervention-induced change in relative abundance of genus *Ruminococcaceae_UCG-003* and intervention-induced protection of stress-induced working memory changes. Subjects with a higher increase in *Ruminococcaceae_UCG-003* abundance after probiotics (right side of the x-axis) were also more protected from negative effects of stress on working memory after probiotics (upper side of the y-axis).

Linear regression was subsequently used to assess potential confounders in the relationship between probiotics-induced changes in working memory performance after stress and *Ruminococcaceae_UCG-003* (Δ scores). Age, BMI at baseline as well as baseline dietary fibre intake were identified as potentially modulating the effect of probiotics and its relation with neurocognitive measures. None of these confounders had a significant effect in the model (model without potential confounders; Standardized Beta *Ruminococcaceae_UCG-003* = 0.45, p=0.018, model with potential confounders; Standardized Beta *Ruminococcaceae_UCG-003* =0.50, p=0.022, potential confounders: Age: Beta=-0.117, p=0.565, BMI: Beta=-0.133, p=0.661, baseline reported fibre intake: Beta=0.170, p=0.814). Dietary intake (DHD-FFQ) scores were not affected by the intervention; all p>0.05 (for scores see **Supplementary Table 1**).

The association between probiotics-induced changes in working memory performance after stress and *Ruminococcaceae_UCG-003* (Δ scores) was not significant in the placebo group (B=-0.179, p=0.296). Indeed, the association was significantly greater for the probiotics’ than for the placebo group: the regression coefficients for the effect of *Ruminococcaceae_UCG-003* on stress-induced working memory changes in the probiotics and placebo groups were significantly different (Slope comparison interaction term: Beta=-0.614, p=0.009).

The exploratory analysis on neural signal during cognitive control and physiological measures did not give significant associations with change in *Ruminococcaceae_UCG-003* abundance, see **Supplementary Results**.

## Discussion

Here, we show that the effects of multi-strain probiotics on stress-related cognitive performance are associated with changes in the gut microbiota composition. This association was exclusive to the probiotics group and independent of the potential confounders age, BMI, and baseline dietary fibre intake.

Healthy, female subjects took a multi-strain probiotics or placebo for 28 days and donated fecal material at baseline and after the intervention period. In an identical pre- and post-intervention session, subjects performed a working memory task before and after an acute stress manipulation. Only those subjects included in the probiotics group showed subtle changes in the beta diversity of their gut microbial communities, but this effect did not significantly differ from the (non-significant) beta diversity change in the placebo group. In terms of gut microbial composition, we observed an increased relative abundance of the genera *Butyricimonas, Alistipes, Christensenellaceae_R-7_group, Family_XIII_AD3011_group, Ruminococcaceae_UCG-003, Ruminococcaceae_UCG-005* and *Ruminococcaceae_UCG-010* in the probiotics versus the placebo group. These nominally increased genera are part of known abundant taxa observed in the healthy human gut ^41,42^. Of these selected genera, only *Ruminococcaceae_UCG-003* was significantly associated with the effect of probiotics on working memory performance after stress, where subjects with a larger probiotics-induced increase in *Ruminococcaceae_UCG-003* abundance also showed a greater protective effect of probiotics on stress-induced working memory changes.

It is the first time that effects of this multi-strain probiotics on gut microbiota composition is tested in humans. In addition to the significant association between *Ruminococcaceae_UCG-003* abundance and stress-related working memory, our results show (uncorrected) probiotics-induced changes in the relative abundance of other genera. Of these, genus *Alistipes, Ruminococcaceae_UCG-005* and *Christensenellaceae* R-7 group have been previously associated with a healthy gut microbial composition when compared with subjects with gastro-intestinal diseases ^43^. Though not all of these genera are classified in function, these three genera as well as *Butyricimonas*, are known to be plant fibre degraders and produce butyrate, a short-chain fatty acid (SCFA) ^44–46^. SCFAs perform a pivotal role in the GBA ^47^, besides being a nutrient source for resident bacteria ^48^. SCFAs beneficially affect the immune system, e.g. by the secretion of cytokines and T-cell differentiation. Moreover, they protect intestinal and blood-brain barrier permeability and modulate the HPA axis through the vagal nerve ^48–52^. Increased SCFA production may hence contribute to the protecting effect of probiotics on gut and brain health.

While we find an association between stress-related cognitive performance and gut microbial abundance, none of the physiological stress measures associated with the gut microbial changes, which is in line with our previous findings that these stress measures were not affected by probiotics across the group ^24^. The stress paradigm did activate the HPA axis as measured by blood pressure, heart rate and cortisol ^24^. Dampened cortisol responses after probiotics’ use is not consistently observed in human RCTs: while Messaoudi et al. ^14^ found lowered urinary levels of cortisol, Takada et al. ^19^ only found lowered levels when pooling salivary cortisol measures from three trials, and Mohammadi et al. ^53^ did not observe serum cortisol changes due to probiotics even though well-being was improved. The absence of physiological effects of the probiotics in the current trial suggests there are other mechanisms at play rather than HPA (de-)activation. For instance, the probiotics’ intervention could have resulted in prevention of stress-induced immune activation that is known to affect brain neurotransmitters, e.g. catecholamines, underlying cognitive performance ^54^. In favor of such a potential mechanism are results using the current multi-species probiotics mixture in rats, where the gut metabolite indolepropionic acid was increased ^16^. Indolepropionic acid is known to improve intestinal barrier function and to limit neuro-inflammation ^55^. SCFAs can also contribute to reduction of stress-induced inflammation ^56,57^. As we did not directly measure gut metabolites in stool or serum, nor inflammatory cytokines, we suggest future studies may look into these functional mechanisms of how probiotics support cognitive resilience to stress.

We were not able to determine if the bacterial strains included in the ‘Ecologic®Barrier’ probiotic product changed after our intervention. The 16S rRNA sequencing technique allows robust and consistent identification of taxa up to the genus category but not lower taxa (i.e. species). None of the genera that were increased by the intervention include any of the supplemented strains. When revising the genera including the strains present in the probiotics mixture, we observed a non-significant increase in relative abundance: genus *Bifidobactium* increased 1,1% post-versus -pre probiotics and genus *Lactobacillus* increased 0.5%. The genus *Lactococcus* was not detected in the sample. It is important to note that the efficacy of probiotics does not depend on and is not expected to be limited to colonization of the supplemented strains themselves. Probiotics can alter relative abundance of multiple bacterial groups, but also affect community dynamics in several ways, e.g. through changes in pH, by outcompeting bacteria by utilizing nutrients, by promoting the growth of resident bacteria through nutrient production, or by occupying adherence sites on the intestinal wall ^48^. In fact, changes in gut microbial composition are not consistently found following probiotics’ use ^48^. Our findings of probiotic-induced changes in other genera than supplemented, therefore, contribute to the understanding of probiotics’ supplementation in healthy human subjects.

Our current findings are in need of replication, ideally in a larger sample and extending the findings to a male sample. Nevertheless, several other studies similarly found variation in *Ruminococcaceae* genera abundance to be associated with neuro-cognitive measures. Even though these findings are not all involving *Ruminococcaceae_UCG-003* specifically, the mentioned genera belong to the *Ruminococcaceae* family, meaning they are evolutionary similar. For example, in a case-control study on Attention Deficit Hyperactivity Disorder (ADHD), *Ruminococcaceae_UCG-003, -004* and *-005* were found more abundant in ADHD patients, and higher levels of *Ruminococcaceae_UCG-004* were associated with a higher number of inattention symptoms (Szopinska-Tokov et al., 2020). Moreover, Bagga et al. ^58^ tested the neuro-cognitive effects of a similar probiotics product in healthy volunteers. They observed that the OTU *Ruminococcaceae_UCG_002* was negatively correlated with depressive feelings. With research into the microbiota-GBA accumulating, it will be interesting to see whether the beneficial role of *Ruminococcaceae* genera in neuro-cognitive performance in health and disease is confirmed.

Studies into the effects of probiotics on neuro-cognition after stress are sparse and even fewer linked these effects to changes in the gut microbiome. In the absence of a stress paradigm, Bagga et al. ^58^ assessed the effect of probiotics on emotional memory and gut microbial composition. Similar to our results, they found that memory performance was improved by the probiotics supplementation, which was in their study associated with changes in the genus *Bacteroides*. This difference in association between the studies, i.e. the currently observed *Ruminococcaceae_UCG-003* vs. *Bacteriodes*, might not be surprising giving the different type of memory (working memory vs. emotional memory), different type of probiotic product (Ecologic®Barrier vs. Ecologic®825), and different sample (larger [n=56 instead of n=23 for microbiota analyses] and more homogenous [only female] in our study). Another study investigated the effect of probiotics in exam-related stress in healthy adults (students) ^20^. In this study, they used a single strain (*Lactobacillus casei* strain Shirota YIT 9029, 1.0 ×10^11 CFU per 100-ml bottle, 8 weeks supplementation; not overlapping with any in the current study). The authors found preservation of within-group diversity after probiotics compared with placebo, whereas in our study between-group diversity shifted in the probiotics group and increased abundance of several genera was observed (on an uncorrected threshold). This difference in functional effects likely partly arises due to the different primer regions of the 16S rRNA gene studied; V1-2 in ^20^ and V4 in our study. These differences in study design (e.g. duration, dose), probiotics product used (e.g. single- or multi-strain), gut microbiome analysis techniques (e.g. in 16S rRNA primer amplicon, DNA extraction method) hamper comparability between probiotics’ studies. Formal replication studies are needed to overcome this limitation to the probiotics field. Accumulation of probiotic trials in the microbiota-GBA field will increase knowledge on the effects, potentially by backtracking shared functional effects (on gut microbial composition, physiology or behavior) to shared properties across these strains.

In conclusion, 4-weeks supplementation of probiotics alters the gut microbial community, nominally increasing the relative abundance of several genera versus placebo. Of these, increased relative abundance of the gut bacterial genus *Ruminococcaccae*_UCG-003 correlates significantly with the positive effects of probiotics on stress-induced working memory changes. In addition to our previous findings of probiotics supporting cognitive performance under stress relative to placebo ^24^, our current results suggest that these beneficial effects are modulated by changes in the gut microbiota community. More research into the functional effects of gut microbial changes would add to the understanding of probiotics’ modulation of the gut-brain axis.

## Data Availability

Data is available upon request.

## Acknowledgements

None

## Conflict of interests

The study was supported by the Dutch Ministry of Economic Affairs under the TKI Life Science and Health, project LSHM15034, and by Winclove Probiotics B.V., The Netherlands. Saskia van Hemert is employed at Winclove Probiotics B.V. This does not alter this authors’ adherence to all publication policies on sharing data and materials. All other authors have no conflicts of interest to declare.

## Notes

### Clinical Trial

NTR5845

### Author Declarations

CMO Arnhem-Nijmegen, NL55406.091.15

