## Supplementary Materials for "Probiotics-induced changes in gut microbial composition and its effects on cognitive performance after stress"

* shared first authors

### shared last authors

Corresponding author: Mirjam Bloemendaal
Address: Radboud university medical center, Genetics Department, Geert Grooteplein 10, 6525 GA Nijmegen, The Netherlands

**Supplementary methods**

*Inclusion criteria*

Subjects were included when using (oral or intra-uterine) hormonal contraceptives and scoring a body mass index (BMI) between 18 and 25. Self-reported medical conditions and regular medication use were exclusion criteria, as well as smoking, lactose intolerance, vegan diet, alcohol intake over ten glasses per week, use of pre- and probiotics supplementation, use of antibiotics within two months, or change in diet three months prior to their participation in the study. Subjects were tested outside of the 'stop week' of oral contraceptives during test sessions to ensure similar hormone levels between both sessions of each participant. For more information on recruitment and drop-out, see [1].

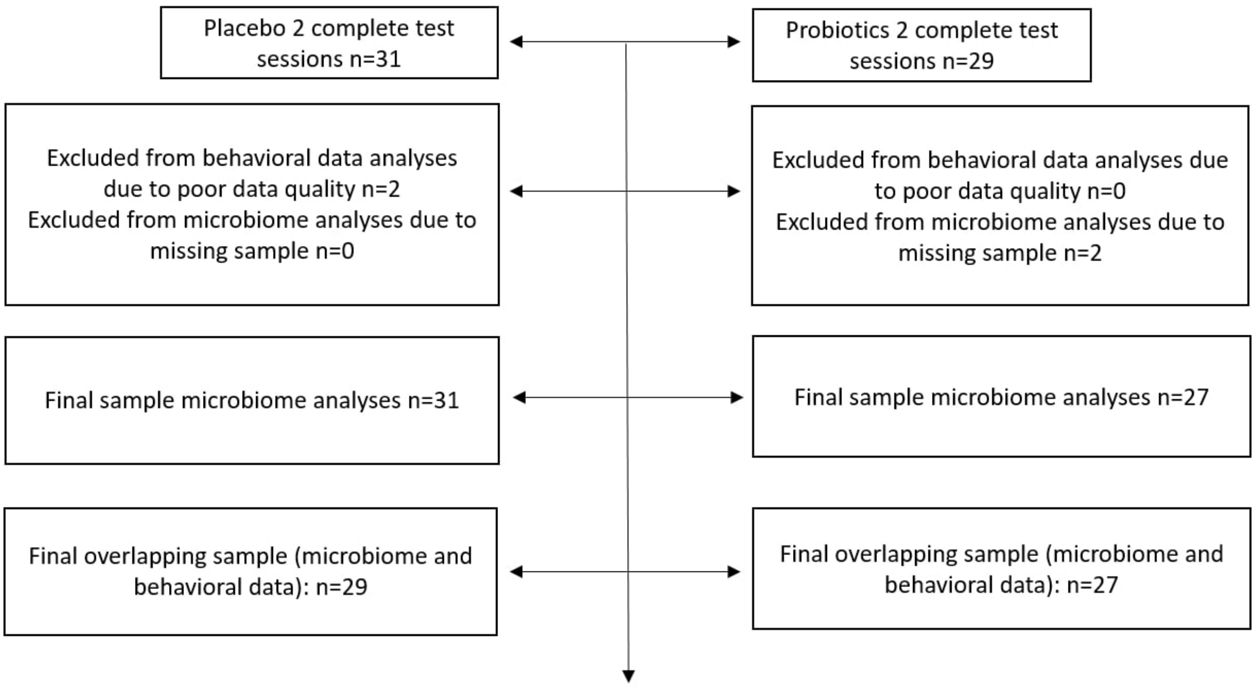


**Supplementary Figure 1.** Consort Flow Diagram

*Intervention*

The probiotic strains were blended into a carrier material consisting of maize starch, maltodextrin, vegetable protein and a mineral mix. The placebo consisted of the same carrier material as used in Ecologic®Barrier and was indistinguishable in color, smell, taste and appearance. Upon completion of the study, subjects answered at chance level whether they had consumed probiotics or placebo product (see [1]). Randomization was performed by Winclove, using a computer-generated scheme in blocks of four. This excluded any involvement of the researchers in the allocation procedure of the subjects.

*Exploratory analyses*

We performed exploratory analyses focused on other relevant measures available. We specifically focused on the measures related to the stress induction, i.e. the physiological stress measures assessed during the SECPT: cortisol, heart rate and blood pressure. Moreover, we tested fMRI signal during the scanner task measuring cognitive control (i.e., the color-word Stroop task), as we found it to be related to the probiotics-induced changes in stress-related working memory in Papalini et al. [1]. We limited our analyses to fMRI signal in three regions of interest (ROIs), i.e. those prefrontal cortex regions that showed a relation with intervention effects on stress-related working memory in the probiotics group (dorsolateral prefrontal cortex, dorsomedial prefrontal cortex and ventrolateral prefrontal cortex, (see Figure 6 in Papalini et al., 2018). For this exploratory analysis, we only took genera correlating with the protective effect of probiotics on stress-induced working memory changes (see **Figure 2 main text**). In three separate repeated measures ANOVA's (within-subjects factor: time, between-subjects factor: intervention, and a covariate of interest: pre- versus post-intervention relative abundance values for genera relating with stress-induced working memory changes), we tested for an interaction term between time, intervention and the covariate. Spearman correlations were performed on fMRI signal (extracted beta values post- minus pre-probiotics) with relative abundance values of the gut microbiota (post- versus pre-probiotics). Significant interactions and correlations were checked for potential confounders and specificity of the effect to the probiotics group (see the approach for association analyses in the main text).

#### Supplementary results

**Supplementary Table 1.** Dutch Healthy Diet Food Frequency Questionnaire (DHD-FFQ) ratings mean (SEM) in the probiotics and placebo group. No baseline differences and no session*group effects observed, all p>0.05).

| DHD-FFQ scales | Probiotics pre-session (N=25) | Probiotics post-session (N=23) | Placebo pre-session (N=27) | Placebo post-session (N=27) |
|---|---|---|---|---|
| Total | 47.45 (2.11) | 51.62 (2.28) | 52.39 (1.79) | 54.49 (2.20) |
| Subscale vegetables | 5.90 (0.50) | 5.85 (0.52) | 6.20 (0.53) | 6.09 (0.54) |
| Subscale fruit | 7.80 (0.45) | 6.80 (0.77) | 7.82 (0.54) | 8.10 (0.49) |
| Subscale fibre | 7.83 (0.41) | 7.82 (0.44) | 7.50 (0.39) | 7.70 (1.76) |
| Subscale fish | 3.96 (0.65) | 4.67 (0.65) | 4.37 (0.52) | 4.48 (3.17) |
| Subscale saturated fat | 3.90 (0.77) | 6.01 (0.88) | 4.10 (0.69) | 5.25 (4.01) |

| | | | | |
|---|---|---|---|---|
| Subscale trans fat | 6.00 (1.00) | 7.39 (0.94) | 8.15 (0.76) | 7.78 (4.01) |
| Subscale salt | 5.96 (0.66) | 6.57 (0.59) | 7.31 (0.41) | 6.79 (2.71) |
| Subscale alcohol | 6.10 (0.81) | 6.52 (0.94) | 6.94 (0.83) | 7.96 (3.54) |

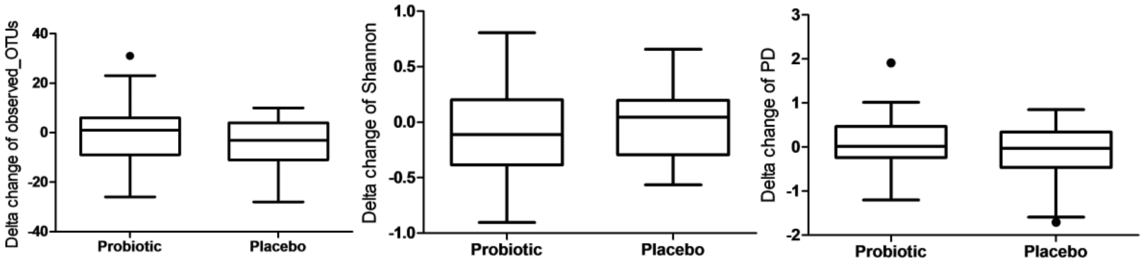


**Supplementary Figure 2.** Alpha diversity measurements of microbial communities in the probiotics and placebo groups. (A) observed OTUs and (B) Shannon and (C) phylogenetic tree (PD).

**Supplementary Table 2.** Beta diversity analysis. PERMANOVA (adonis) results on weighted UniFrac dissimilarity matrix. Significant p-values are marked in bold.

| Variables | N | R2 | Pseudo-F | P-value |
|---|---|---|---|---|
| Time (Pre, Post) * Intervention (Placebo, Probiotics) | 116 | 0.003 | 0.36 | 0.244 |
| Main effect of Time (Pre vs. Post) | 116 | 0.010 | 1.20 | **0.011** |
| Main effect of Intervention (Placebo vs Probiotics) | 116 | 0.003 | 0.36 | 0.703 |
| Placebo group Pre vs. Post | 62 | 0.006 | 0.39 | 0.210 |
| Probiotics group Pre vs. Post | 54 | 0.024 | 1.27 | **0.008** |
| Pre vs. Pre | 58 | 0.005 | 0.31 | 0.754 |
| Post vs. Post | 58 | 0.008 | 0.42 | 0.655 |

Note: significant p-values in are indicated in bold.

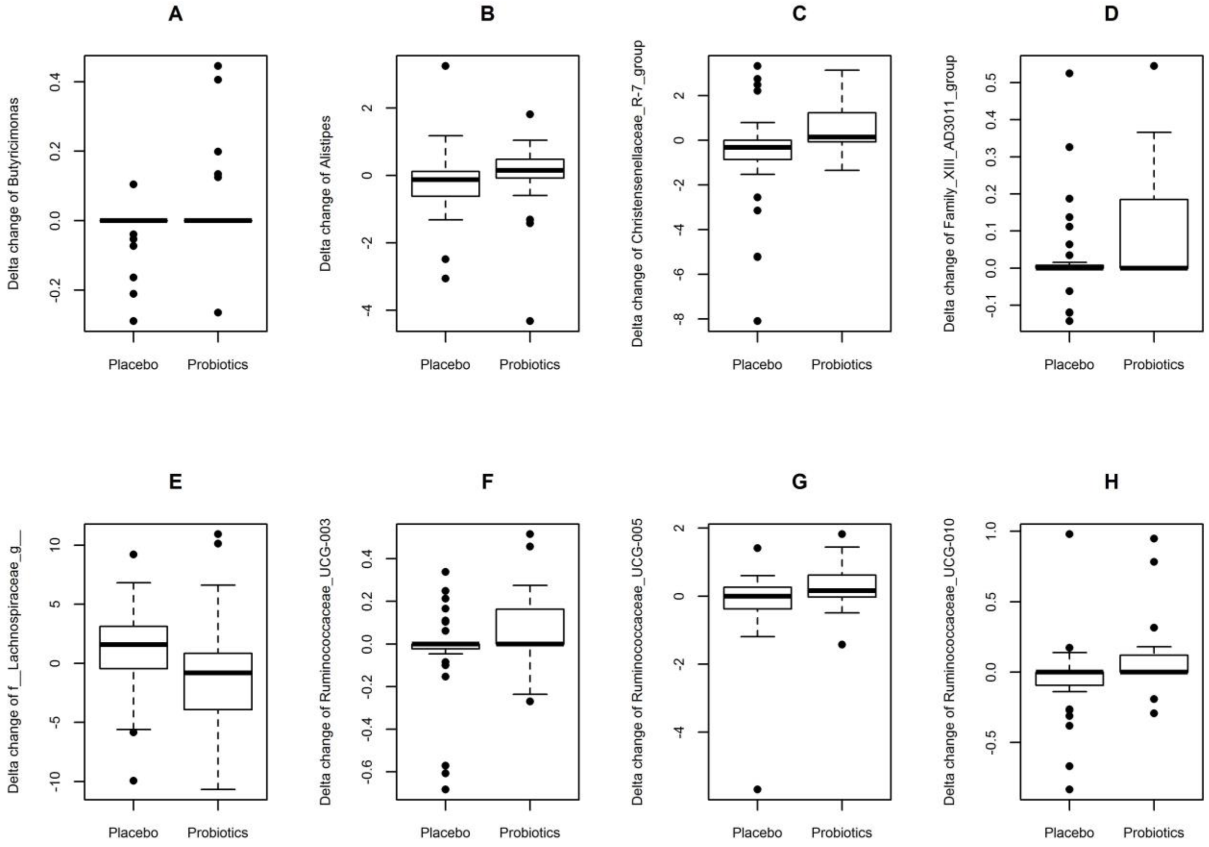


**Supplementary Figure 3**. Boxplot representing changes of genera that differ significantly between Δ probiotics and Δ placebo groups. (A) Butyricimonas, (B) Alistipes, (C) Christensenellaceae_R-7_group, (D) Family_XIII_AD3011_group, (E) f_Lachnospiraceae;g_, (F) Ruminococcaceae_UCG-003, (G) Ruminococcaceae_UCG-005 and (H) Ruminococcaceae_UCG-010.

**Supplementary Table 3.** Comparison of phylum relative abundance post-pre intervention changes (Δ) between probiotics and placebo groups.

| | ΔProbiotics Mean Rank | ΔProbiotics Mean (range) | ΔPlacebo Mean Rank | ΔPlacebo Mean (range) | P-value* |
|---|---|---|---|---|---|
| Euryarchaeota | 31.89 | 0.0005 (-0.015- 0.014) | 27.42 | -0.0015(-0.021-0.011) | 0.211 |
| Actinobacteria | 30.19 | 0.0117 (-0.166-0.351) | 28.90 | -0.0089(-0.270-0.176) | 0.773 |
| Cyanobacteria | 33.41 | 0.0024 (-0.001-0.038) | 26.10 | 0.0004(-0.010-0.020) | 0.063 |
| Proteobacteria | 33.52 | -0.0004 (-0.020-0.011) | 26.00 | -0.0059 (-0.040-0.021) | 0.091 |
| Tenericutes | 29.85 | 0.0001 (-0.005-0.010) | 29.19 | -0.0011 (-0.027-0.004) | 0.863 |
| Verrucomicrobia | 32.89 | -0.0053 (-0.126-0.059) | 26.55 | -0.0013 (-0.092-0.173) | 0.151 |
| Bacteroidetes | 29.78 | -0.0322(-0.242-0.142) | 29.26 | -0.0365(-0.329-0.265) | 0.907 |
| Firmicutes | 27.22 | 0.0230 (-0.155- 0.198) | 31.48 | 0.0547 (-0.244-0.372) | 0.338 |

* P-values (uncorrected) are given for non-parametric Mann-Whitney U test assessing the differences in rank order. These non-parametric rank tests were performed due to the overall skewness of the microbiota data including many zero values. A cut-off was placed including maximally 90% zero values (see the Method section for more details).

*Exploratory analyses*

Our exploratory analyses, using the *Ruminococcaceae_UCG-003* genus showed no interaction between the increase in relative abundance of this genus and probiotics' induced changes in cortisol, heart rate or blood pressure (p >.05). Linear regression on the probiotics' effect on neural signal in the frontal ROIs activated during the cognitive control (i.e., Stroop) task (Δ scores of dorsolateral prefrontal cortex, dorsomedial prefrontal cortex and ventrolateral prefrontal cortex ROIs, see Papalini et al., 2018) showed that these neural signals were not associated with the probiotics-induced increase in the relative abundance of the *Ruminococcaceae_UCG-003* genus (all regression beta's p > 0.05).
